# Beyond Image Recognition: Applying Deep Learning to List-Type Medical Data for Risk Prediction

**DOI:** 10.1101/2024.10.31.24316486

**Authors:** Gaku Kondo

## Abstract

**Objective:** The purpose of our research was to develop efficient utilization of Deep Learning to the list-type data in which medical characteristics are arranged.

**Materials and Methods:** We conducted a survey of blood donors, focusing on the rare adverse reaction of falling. In addition to all cases of fainting, we randomly selected a control group of donors who did not fall. This data was then converted into a two-dimensional format suitable for analysis with a convolutional neural network (CNN). We used an under sampling method to randomly divide the dataset into training and testing sets. Finally, we used the CNN and a logistic regression model to predict the probability of fainting, calculate anomaly scores, and rank the risk of falling.

**Results:** The convolutional neural network (CNN) identified 3 out of 10 falls among the group with the top 1% of anomaly scores. In contrast, the logistic regression model failed to identify any falls within the same top 1% anomaly score group.

**Conclusion:** Applying information converted into two-dimensional data to Deep Learning by using anomaly detection together was useful to narrow down with high-risk group. Although these findings require validation in a larger and more diverse population, the success of this approach in predicting falls after blood donation suggests its potential for predicting other rare adverse events in healthcare, such as adverse drug reactions, complications from medical procedures, or even disease outbreaks.

## BACKGROUND

Currently, Artificial Intelligence (AI) is attracting attention. In particular, the use of Deep Learning is remarkable and has been successful in many fields such as image recognition used in automatic driving systems and voice recognition in smartphones.^1^ As for the application of Deep Learning in the medical field, it demonstrates excellent performance for image analysis such as Computed Tomography (CT) and Magnetic Resonance Imaging (MRI). In recent years, it has also been used for electrocardiogram (ECG) analysis.^2^ On the other hand, there is currently no great progress in application to list-type data in which medical characteristics are arranged in one line. This time, we applied Deep Learning to the list-type data, which was converted into two-dimensional data, for the purpose of estimating the risk of occurrence of a rare adverse reaction after collecting blood, so we report the details.

## METHODS

### Study design and population

This is a retrospective cross-sectional study, conducted by extracting the records of blood donors stored in the database server of the Japanese Red Cross Society. The ethics committee of the Japanese Red Cross Society, chaired by Shuichi Kino, approved this study (Ethical review number: 2024-016), and the consent to study participation was obtained in the form of opt-out on the webpage. In this analysis, we collected those who actually made a whole blood donation of 400 mL in Tokyo between January 1 and December 31, 2019. Considering the peculiarity that the coronavirus disease 2019 pandemic may impart to the collected data, we used the sample in the pre-pandemic period.

### Data collection

A survey of the study population identified a total of 32 reports of falls among 361,114 donors, which may not be all cases, since falls outside the venue were entered into the database only when contacted. Those who donated blood multiple times during this period were treated as separate individuals. In addition to the 32 falls, 100,210 samples were randomly selected from the whole donors of 361,082 who did not fall during the study period. In addition to the presence or absence of falls, the extracted 100,242 data the number of past vasovagal reaction occurrences, blood donation history (first-time or repeated), sex, height, weight, and Body Mass Index (BMI), pulse pressure divided by systolic blood pressure (PP/sBP), shock index (pulse rate divided by systolic blood pressure), age, sleep time (sleep time in the previous night), fasting time (the elapsed time since the last meal), and the blood donation venue (mobile or fixed). These variables were arranged in this order in each one-dimensional vector. Note that the vital signs were the values before blood collection.

### Outcome

The objective variable is the occurrence (positive/negative) of fall after blood collection. The object of evaluation is the accuracy for the estimated risk of falling. Fall occurs as a result of vasovagal syncope caused by the blood loss, with a frequency of approximately 0.01% of all blood donors in Tokyo. There are many accidental factors about the development, so it is extremely difficult to predict in advance.

### Deep Learning

A neural network is understood as a network of units (artificial neurons) modeled neurons in the human brain. The units combine input values, perform computations and output new values. Deep Learning is the layering of this network deep. There are several types of deep learning. In this analysis, we used Convolutional Neural Network (CNN), which has been in the limelight in recent years. CNN is modeled on human vision and made up of multiple layers consisting of convolutional layers and pooling layers, which is not just a “deep layer” but also a structure. Two-dimensional data is scanned while a sheet called a filter is moved, and the results are recorded in the convolutional layer units. The results of the scan recorded in the convolutional layer are further bundled and concentrated into units in the pooling layer.^3,4^ As a result, CNN can efficiently deal with complex pattern recognition problems.

### Data analysis

Our 100,242 sample to be analyzed was an imbalanced data with remarkably skewed class proportions for the objective variable (positive case: 32 vs. negative case: 100,210). To learn efficiently on the imbalanced data, data splitting with some ingenuity was required. As training data, 22 out of 32 fall cases and 220 out of 100,210 non-fall cases were randomly selected, respectively. The remaining 100,000 cases were used as test data. The sample size (100,240 extracted from the whole) was set so that the falling rate of the test data was 1/10,000. Such a division method is called under-sampling and is often used in the field of machine learning as a countermeasure against imbalanced data.

After we trained CNN model and Logistic Regression (LR) representing conventional statistics to learn the patterns of fall occurrence in the training data, we predicted the probability of falling for individuals in the test data using the two learning models. Next, we verified the discriminative ability of the two predictive models using the method of anomaly detection on the predicted probability. Anomaly detection is a technique for detecting samples that behave differently compared to the majority of the accumulated data. ^5^ We performed anomaly detection based on gamma distribution, in which the two parameters, Shape and Scale, were estimated by the method of moments.^5^ Anomaly score was calculated as the minus logarithm of the probability density in the gamma distribution for each sample.^5^ The variables used for the learning were as follows. The metric scale variables were age, height, weight, BMI, PP/sBP, shock index measured before blood collecting, sleep time, fasting time, number of past vasovagal reaction occurrences. The categorical scale variables were sex, blood donation history (first-time or repeated), and the blood donation venue (mobile or fixed). Categorical scale variables were converted to numerical data of 0 and 1 as dummy variables. As data preprocessing, age, weight, and BMI were divided by 10, and shock index and PP/sBP was multiplied by 10 to align the number of digits between variables.

Since CNN uses two-dimensional data (matrix) as learning objects, the list-type data of row vectors consisting of 12 variables had to be converted to 12 × 12 matrices. We took the geometric mean between each variable as the upper triangular element of the matrix, imitating a round-robin combination table. arithmetic mean between each variable were substituted for the lower triangular elements of the matrix. As a result, the original elements of the row vector were located over the diagonal of the matrix. Each average was rounded to the nearest whole number. A specific example is shown in Figure 1.

**Figure 1.**
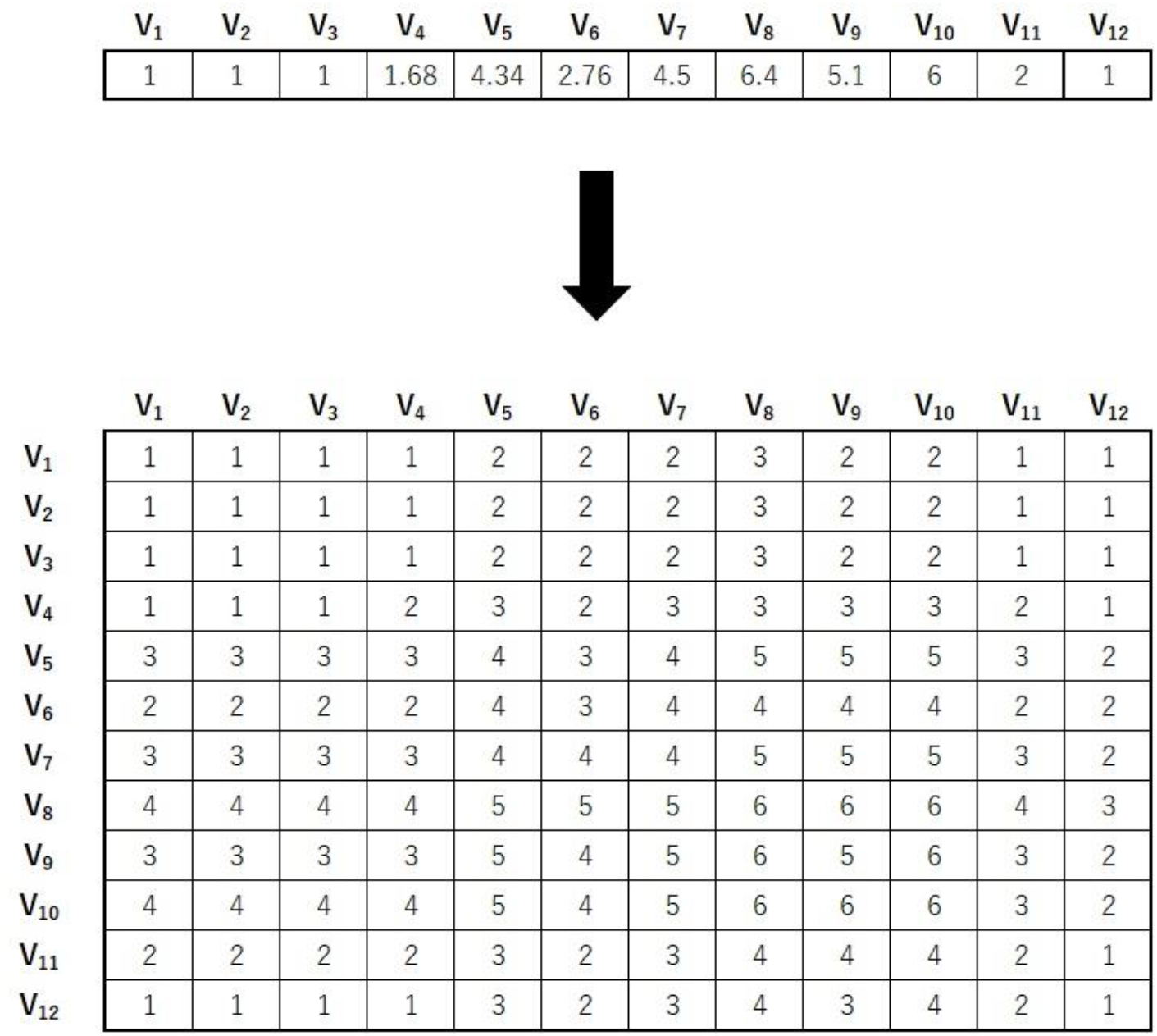
An example of conversion from a row vector of 12 elements to a 12 × 12 matrix. The upper triangular part of the matrix consists of the geometric mean of the values of two variables. The element of the i-th row and the j-th column is the geometric mean of the two values of Vi and Vj (i ≤ j). The lower triangular part consists of the arithmetic mean of the values of two variables. The element of the i-th row and the j-th column is the arithmetic mean of the two values of Vi and Vj (i > j). Each average was rounded to the nearest whole number. Vk is an abbreviation for the k-th variable.

The CNN model we constructed in learning data consisted of two layers of convolutional layer and pooling layer. Although detailed settings are omitted, the data matrix of size 12 was reduced to size 5 by the first convolution/pooling and was reduced to size 2 by the second convolution/pooling. We used 64 different filters, resulting in 64 data matrices of size 2. The probability of belonging to each class was calculated by regressing 2 × 2 × 64 = 256 values compressed in this way ^4^. The parameters were initially set based on empirical rules, as was the selection of functions used during the learning process, such as the activation and loss functions. The filter size and number of paddings were then tuned using the hold-out method.

The hyperparameters for the model were set as follows: the activation function was a rectified linear function, the loss function was a SoftMax cross entropy, and the pooling method was Max Pooling with a size of 2×2. The model was trained for 10 epochs. In the first convolutional layer, the filter size was 5×5 with a stride of 1 and padding of 1. For the second convolutional layer, the filter size was 2×2 with a stride of 1 and no padding. A framework called Chainer (ver.4.0.0) was used to perform CNN Learning. Chainer is a Python-based Deep Learning framework published by Preferred Networks.^7^ We used scikit-learn (version 0.24.1), a Python machine learning library, to perform logistic regression. Our analysis also utilized other Python libraries, such as NumPy and Matplotlib.

## RESULTS

### Population’s characteristics

Blood donor characteristics in our extracted data are shown in Table 1.

**Table 1.**
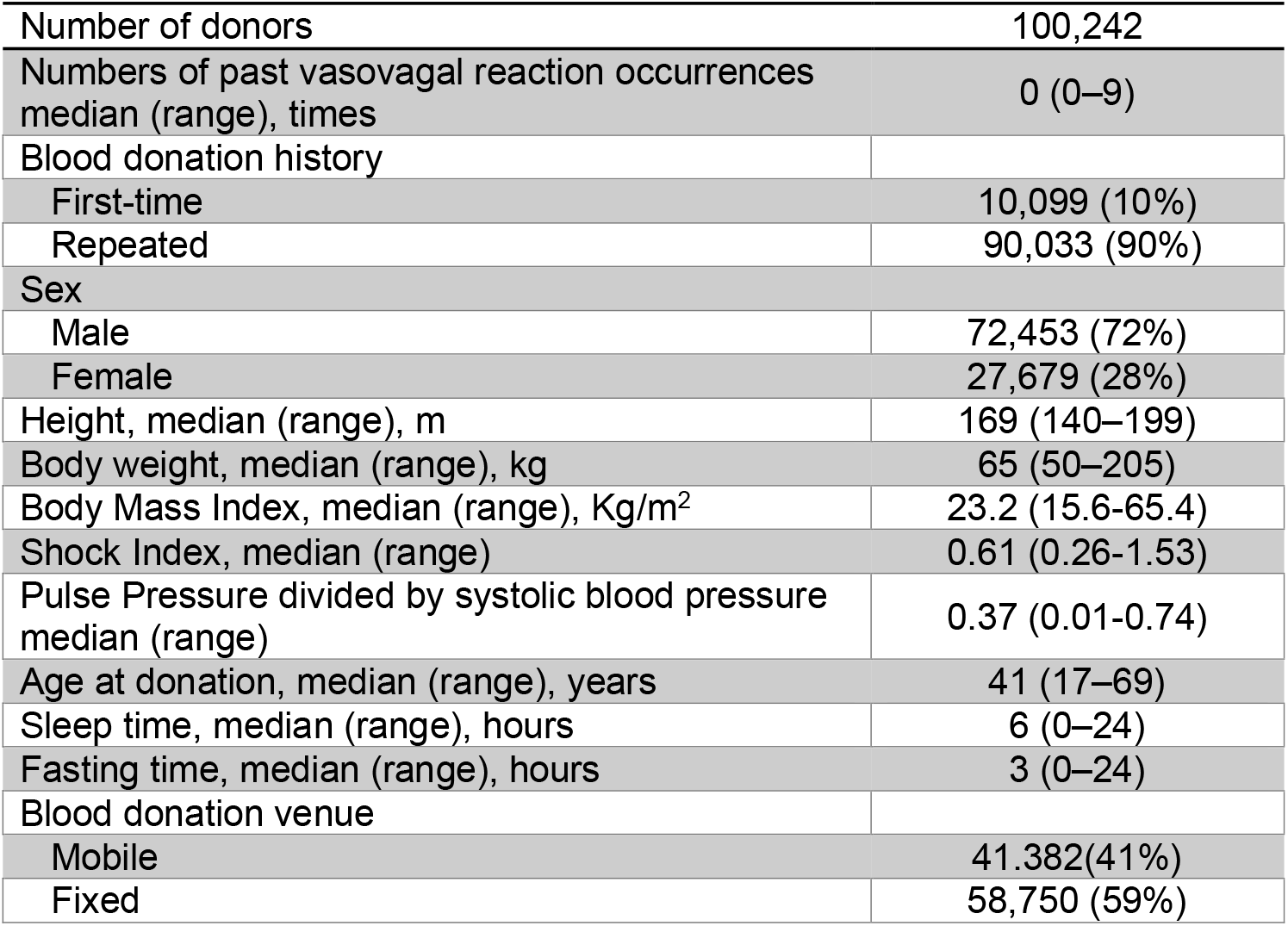
Population’s characteristics in our extracted data.

|  |  |
| --- | --- |
| Number of donors | 100,242 |
| Numbers of past vasovagal reaction occurrences<br>median (range), times | 0 (0–9) |
| Blood donation history |  |
| First-time | 10,099 (10%) |
| Repeated | 90,033 (90%) |
| Sex |  |
| Male | 72,453 (72%) |
| Female | 27,679 (28%) |
| Height, median (range), m | 169 (140–199) |
| Body weight, median (range), kg | 65 (50–205) |
| Body Mass Index, median (range), Kg/m<sup>2</sup> | 23.2 (15.6-65.4) |
| Shock Index, median (range) | 0.61 (0.26-1.53) |
| Pulse Pressure divided by systolic blood pressure<br>median (range) | 0.37 (0.01-0.74) |
| Age at donation, median (range), years | 41 (17–69) |
| Sleep time, median (range), hours | 6 (0–24) |
| Fasting time, median (range), hours | 3 (0–24) |
| Blood donation venue |  |
| Mobile | 41,382(41%) |
| Fixed | 58,750 (59%) |

### Outcomes

As a result of calculating the anomaly score on the predicted possibility in CNN Learning based on gamma distribution, the summary had a median of 1.12 and a range of 0.99–7.78, and the anomaly score in the top 1% was 4.8. Using this value as a guideline, the threshold for anomaly detection was set to 4.8. Figure 2 shows the distribution of the anomaly scores for the predicted possibilities. As a result of anomaly detection with a threshold of 4.8, 961 anomaly samples were detected, including 3 falls (Table 2A). For the anomaly score on the predicted possibility in LR Learning based on gamma distribution, the summary had a median of –1.16 and a range of –5.93–6.55, and the anomaly score in the top 1% was 4.6. Using this value as a guideline, the threshold for anomaly detection was set to 4.6. As a result of anomaly detection with a threshold of 4.6, 1,040 anomaly samples were detected, including 0 falls (Table 2B).

**Table 2.** The result of anomaly detection with a threshold equal to the top 1%.

**A CNN Learning**
|  | **Case of falling** | **Total number** | **Percentage of falling** |
| --- | --- | --- | --- |
| **Group judged anomaly** | 3 | 961 | 0.31% |
| **Group judged non-anomaly** | 7 | 99,039 | 0.0071% |

**B LR Learning**
|  | **Case of falling** | **Total number** | **Percentage of falling** |
| --- | --- | --- | --- |
| **Group judged anomaly** | 0 | 1,040 | 0% |
| **Group judged non-anomaly** | 10 | 98,960 | 0.01% |

**Figure 2.**
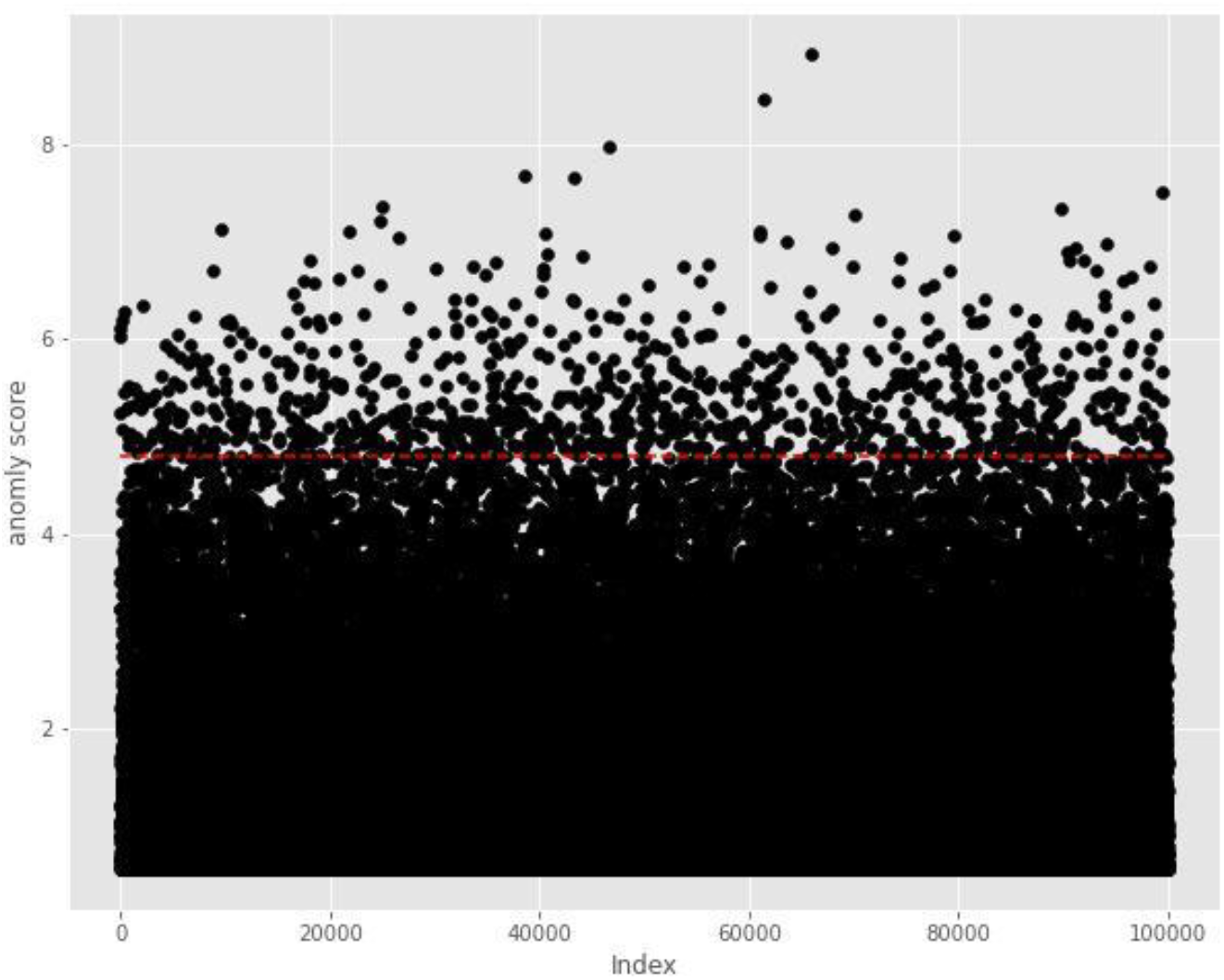
Anomaly detection based on gamma distribution in CNN model, showing the distribution of the anomaly scores for the predicted possibility in the test data. Anomaly score was calculated as the minus logarithm of the probability density in the gamma distribution for each sample in the test data. The red dashed line is the threshold, using an anomaly score of 4.8, above which the sample was considered abnormal.

## DISCUSSION

As a result of calculating the anomaly score using the gamma distribution on the predicted probability calculated by CNN for the test data, among the 100,000 samples, the top 1% samples had 3 falls, and the risk ratio of falling in the group judged anomaly to the group judged non-anomaly was 44.2. On the other hand, by LR Learning, the top 1% samples had 0 falls, and the risk ratio of falling in the group judged anomaly to the group judged non-anomaly was 0. A fall after blood donation is an event expressed as “extreme event,” which has happened very rarely, and standard statistical methods are understood to be inapplicable. We think that the risk assessment by CNN may be a promising approach for such an extreme event. We used the geometric and arithmetic mean between each variable when converting a horizontal one-column vector consisting of the subject’s attributes and measured values into a matrix that is the learning target of the CNN. This is just one attempt, and there may be more effective conversion methods.

This study has some limitations. First, the number of falls, which was the objective variable, was too small. In such a situation, it is presumed that the variation in verification results will increase. In addition, the hold-out method, which divides the data into the two sets, was used for the internal verification of the predictive model, but it has been reported that this method also results in large variations in the verification results.^8^ Information may be lacking on the versatility of the results. Second, due to the severe imbalance of data, our study was forced to be a retrospective study, which generally produces lower-quality evidence compared to prospective study.

This approach has important implications for preventive medicine and public health. By identifying individuals at high risk of falling, targeted interventions can be implemented to mitigate that risk. This could include closer monitoring after donation, providing personalized advice on pre-donation preparation (e.g., hydration, food intake), or even deferring donation for those deemed at highest risk.

By reducing the incidence of falls and subsequent complications, we improve the efficiency of blood collection, and resources can be used more effectively. Although these findings require validation in a larger and more diverse population, the success of this approach in predicting falls after blood donation suggests its potential for predicting other rare adverse events in healthcare, such as adverse drug reactions, complications from medical procedures, or even disease outbreaks.

## CONCLUSION

Applying information before blood collection converted into two-dimensional data to Deep Learning by using anomaly detection together was useful to narrow down with high risk of falling. This method could lead to the improvement of the risk assessment in clinical practice that has been difficult to evaluate with conventional methods.

## Data Availability

All data produced in the present study could be available upon reasonable request to the authors, subject to the consent of the Japanese Red Cross Society.

